# Deep learning-based end-to-end automated stenosis classification and localization on catheter coronary angiography

**DOI:** 10.1101/2021.03.05.21252217

**Authors:** Chao Cong, Yoko Kato, Henrique D. Vasconcellos, Mohammad R. Ostovaneh, Joao A.C. Lima, Bharath Ambale-Venkatesh

## Abstract

**Background:** Automatic coronary angiography (CAG) assessment may help in faster screening and diagnosis of patients. Current CNN-based vessel-segmentation suffers from sampling imbalance, candidate frame selection, and overfitting; few have shown adequate performance for CAG stenosis classification. We aimed to provide an end-to-end workflow that may solve these problems.

**Methods:** A deep learning-based end-to-end workflow was employed as follows: 1) Candidate frame selection from CAG videograms with CNN+LSTM network, 2) Stenosis classification with Inception-v3 using 2 or 3 categories (<25%, >25%, and/or total occlusion) with and without redundancy training, and 3) Stenosis localization with two methods of class activation map (CAM) and anchor-based feature pyramid network (FPN). Overall 13744 frames from 230 studies were used for the stenosis classification training and 4-fold cross-validation for image-, artery-, and per-patient-level. For the stenosis localization training and 4-fold cross-validation, 690 images with >25% stenosis were used.

**Results:** Our model achieved an accuracy of 0.85, sensitivity of 0.96, and AUC of 0.86 in per-patient level stenosis classification. Redundancy training was effective to improve classification performance. Stenosis position localization was adequate with better quantitative results in anchor-based FPN model, achieving global-sensitivity for LCA and RCA of 0.68 and 0.70 with mean square error (MSE) values of 39.3 and 37.6 pixels respectively, in the 520 × 520 pixel image.

**Conclusion:** A fully-automatic end-to-end deep learning-based workflow that eliminates the vessel extraction and segmentation step was feasible in coronary artery stenosis classification and localization on CAG images.

**Key Points:**

1. The fully-automatic, end-to-end workflow which eliminated the vessel extraction and segmentation step for supervised-learning was feasible in the stenosis classification on CAG images, achieving an accuracy of 0.85, sensitivity of 0.96, and AUC of 0.86 in per-patient level.
2. The redundancy training improved the AUC values, accuracy, F1-score, and kappa score of the stenosis classification.
3. Stenosis position localization was assessed in two methods of CAM-based and anchor-based models, which performance was acceptable with better quantitative results in anchor-based models.

**Summary Statement:** A fully-automatic end-to-end deep learning-based workflow which eliminated the vessel extraction and segmentation step was feasible in the stenosis classification and localization on CAG images. The redundancy training improved the stenosis classification performance.

## Introduction

Coronary artery disease (CAD) is the leading cause of morbidity and mortality worldwide [1]. X-ray coronary angiography (CAG) is the current gold standard imaging technique for CAD diagnosis. Expert CAG interpretation requires considerable ‘hands on’ training both visually and cognitively. In clinical practice and also for quality control purposes, screening CAG studies visually to distinguish cases with normal or mild stenosis from those with clinically significant stenosis is a time-consuming process even for experienced readers. Developing an automatic CAG assessment tool to exclude normal or mild stenosis cases would facilitate diagnosis and treatment and enable the screening of large data sets for quality control purposes.

The current “automated or semi-automated” CAG stenosis detection method consists of multiple steps. For example, the most widely used vessel-based workflow starts from the visual or automatic selection of candidate frames from a CAG video [2]. This is followed by the artery extraction using image segmentation algorithms [3] like center-tracking [4]–[6], model-based [7], [8], or Convolutional Neural Network (CNN) [9]–[12]. Finally, individual stenotic lesion localization and classification is performed [4], [5], [8]. The need for extensive human interaction during image data and training label preparation, in addition to addressing problems of sampling imbalance during supervised-learning, has led to algorithms that are commonly evaluated on small datasets prone to overfitting. In this study, we propose an end-to-end workflow for (a) easier data preparation and labeling of large datasets, (b) ruling out significant stenosis, and (c) allowing for precise localization of coronary stenoses in CAG images.

## Methods

### Study population

This research was retrospectively performed on 230 participants with available data from a “Combined Non-invasive Coronary Angiography and Myocardial Perfusion Imaging Using 320 Detector Computed Tomography (CORE320)” study (www.clinicaltrials.gov, NCT00934037), a prospective, multicenter, international study that assessed the performance of combined 320-row CTA and myocardial CT perfusion imaging (CTP) in comparison with the combination of invasive coronary angiography and single-photon emission computed tomography myocardial perfusion imaging (SPECT-MPI) for detecting myocardial perfusion defects and luminal stenosis in patients with suspected coronary artery disease [13][14]. For the stenosis classification, thirty-six studies out of 230 were excluded from the training due to the low image quality or contrasting condition. These images however, were included for evaluation. The original CORE320 study was approved by central and local institutional review boards, and written informed consent was obtained from all participants[13][14]. Given the retrospective and ancillary nature of the data, the current study is covered by the original CORE320 study IRB.

### Candidate Frame Selection

The entire study workflow is summarized in Figure 1. All the CAG studies were saved in the universal DICOM format with a resolution of 512 × 512, 15 fps, typically 60-200 frames per view. Coronary type (left and right coronary artery, LCA and RCA) was classified initially by experts in a small subset (19 patients). This was then leveraged by training a simple classifier [15] for automated coronary selection (100% classification accuracy was obtained). To identify the angle views of the CAG images, DICOM tags were used. Overall 4 angles for LCA (LAO Cranial, LAO Caudal, RAO Cranial, and RAO Caudal) and 3 angles for RCA (LAO, straight RAO, and shallow LAO/RAO Cranial) were used based on the optimal view map (OVM) [16]. A candidate frame was defined as an image with good quality, full contrasting, clear vessel border, and anatomical significance of stenosis (if it had stenosis) in a video frame. A redundancy frame was defined as a background frame without any contrasting agent in arteries. Redundancy frames were added to the training dataset but not in the validation. Exploiting redundancy to improve classification accuracy has been used before [17]. Each training category was provided with the same prevalence of candidate frame and redundancy frame to avoid category imbalance.

**Figure 1.**
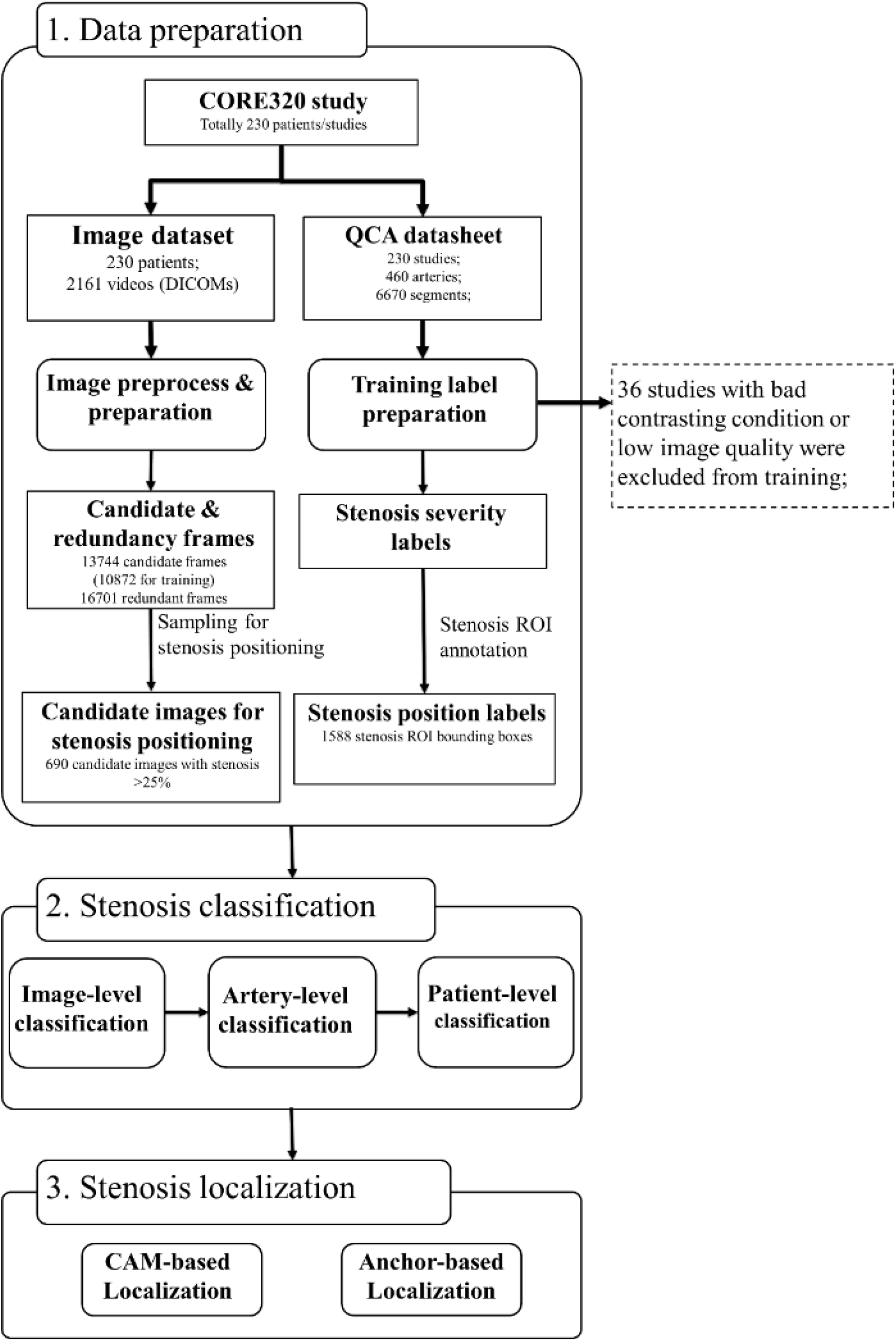
Dataset and Algorithm Workflow. Three steps of data preparation, stenosis classification, and stenosis positioning were presented. The steps of image and training label preparation including coronary artery selection, viewing angle selection, and contrasting frame detection was designed in a fully automatic manner. Stenosis severity classification training was performed in image-level, artery-level, and patient-level. Stenosis positioning was performed in two methods of CAM-based and anchor-based methods. CAM = class activation map. QCA = quantitative coronary angiography.

A CNN+LSTM network was implemented for the candidate frame selection from 19 patients (146 videos in total, and 18688 frames overall). Inception-v3 was employed as a basic classifier to recognize full-contrasting frames and non-contrasting frames as candidates or redundancy frames. Then, the fully-connection layer was connected to a bi-directional LSTM. The inception model was pre-trained for 200 epochs with the initial learning rate (LR) of 1e-4 and the loss function as binary entropy. Then LSTM was trained for 100 epochs with LR=4e-5 and the loss function was defined as convolutional F1 score. Typically, this strategy selected 5 to 10 candidate frames per video.

The performance of candidate frame detection was tested with 582 videos from 175 patients using mean error and standard deviations of beginning contrasting frame (BCF) and ending contrasting frame (ECF) between ground-truth and prediction. The acceptance and error rates were also calculated with average differences of BCF and ECF in a pre-defined range [2], in which accept rate with the error <=3 and error rate with the error >=10. Performance was reported using classification accuracy, F1 and Kappa.

### Stenosis Classification

QCA results previously documented in the CORE320 study were utilized, which documented the 29-segment model localization [18] with their per-segmental stenosis severities. For our study goals (ruling out significant stenosis), coronary stenosis severities were re-categorized into three groups of <25% stenosis, 25 to 99% stenosis, total occlusion as 3-CAT, and two groups of <25% stenosis, ≥25% stenosis as 2-CAT. These were used for artery-level labels. For the image-level labeling, per-segmental stenosis categories were assigned in each angle-view.

Inception-v3 was employed for the stenosis classification network with and without additional redundancy frames in the training dataset to compare the performance with redundancy. The stenosis classification training was performed on 4 models of LCA for each angle view and one model of RCA combining the three angle views due to the complicated features of LCA when compared to the RCA. A max-pooling layer was added to the output of inception to evaluate the artery-level stenosis prediction and for the patient-level stenosis prediction. (Figure 1, 2).

**Figure 2.**
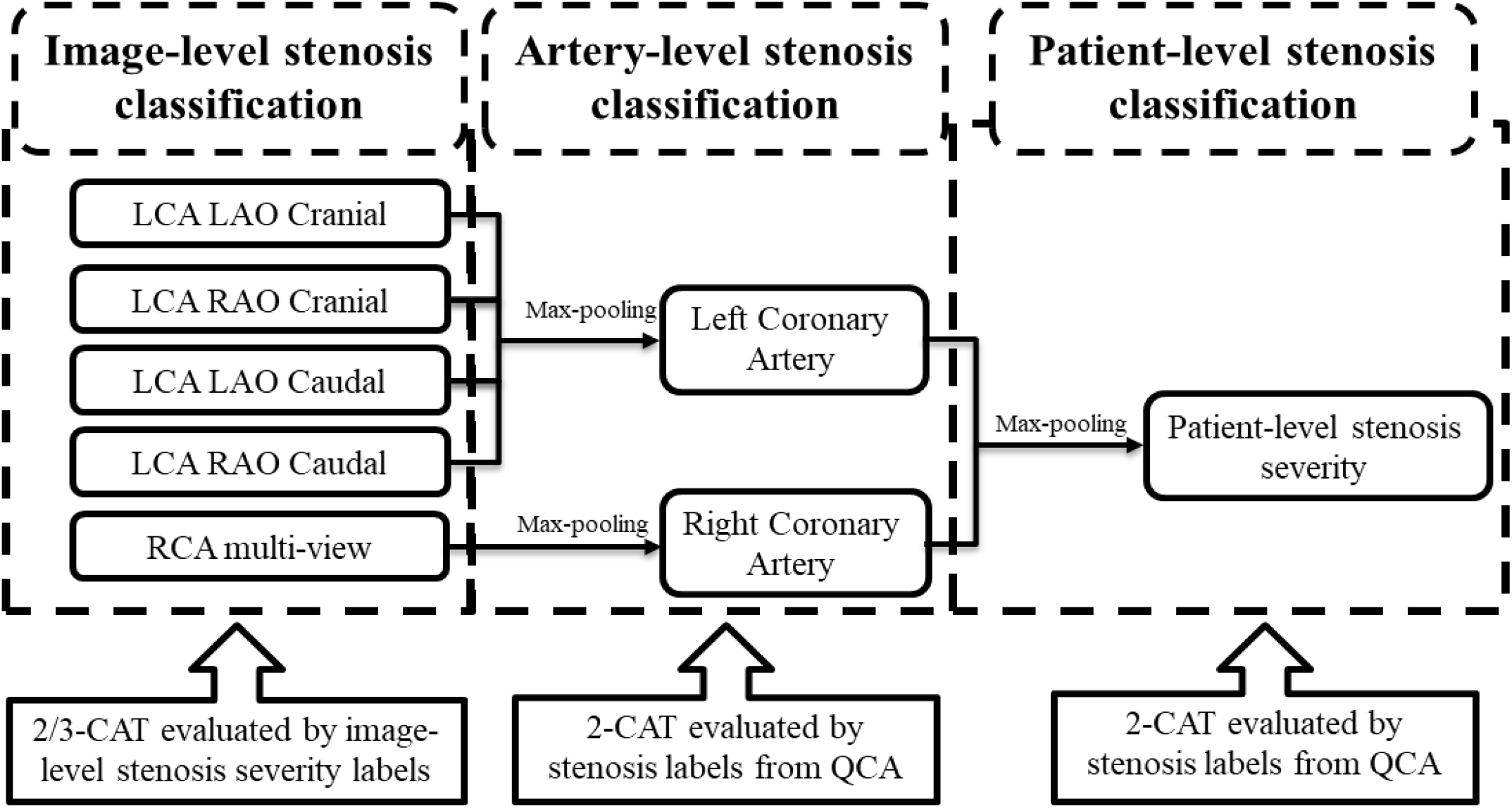
The architecture of the output of the stenosis classification inception model. A max-pooling layer was added to the output of inception to evaluate the artery-level stenosis prediction and for the patient-level stenosis prediction. LCA = left coronary artery. LAO = left anterior oblique. RAO = right anterior oblique. RCA = right coronary artery. QCA = quantitative coronary angiography.

Overall 10872 frames from 194 studies were used for image-level stenosis classification training and 13744 frames from 230 studies were used for the 4-fold cross-validation. Distribution of the cases in the image-, artery-, and patient-levels are summarized in Table 1. The performance of image-level stenosis classification results of LCA was reported as a combined result of four angles. Performance of image-level classification on 3-CAT and 2-CAT with and without redundancy training was reported using accuracy, sensitivity, F1, Kappa, and area under the curve (AUC). Performance of artery-level and per-patient level classification was assessed on the 2-CAT with redundancy training image-level results and reported using accuracy, sensitivity, and AUC.

**Table 1.** Distribution of the cases in each stenosis severity categories used for the image view, artery, and patient-levels validation.

| Stenosis severity | Image level<br>(LCA-LAO Cranial) | Image level<br>(LCA - RAO Cranial) | Image level<br>(LCA – LAO Caudal) | Image level<br>(LCA – RAO Caudal) | Image level<br>(RCA) | Coronary artery level<br>(LCA) | Coronary artery level<br>(RCA) | Per-patient level |
| --- | --- | --- | --- | --- | --- | --- | --- | --- |
| <25% | 624<br>(32.7%) | 752<br>(33.0%) | 673<br>(43.8%) | 789<br>(34.7%) | 2292<br>(39.9%) | 57<br>(24.8%) | 84<br>(36.5%) | 46<br>(20.0%) |
| 25% to 99% | 1123<br>(58.9%) | 1393<br>(61.1%) | 673<br>(43.8%) | 1333<br>(58.5%) | 3181<br>(55.4%) | 132<br>(57.4%) | 118<br>(51.3%) | 127<br>(55.2%) |
| 100% | 160<br>(8.4%) | 135<br>(6.0%) | 189<br>(12.3%) | 155<br>(6.8%) | 272<br>(4.7%) | 41<br>(17.8%) | 28<br>(12.2%) | 57<br>(24.8%) |

Overall 13744 frames from 230 studies were used for the image level validation. Each of the coronary artery level and per-patient level validations were performed on 230 cases. LCA = left coronary artery. LAO = left anterior oblique. RAO = right anterior oblique. RCA = right coronary artery.

### Stenosis Localization

For the stenosis positioning, two methods were investigated: 1) class activation map (CAM) [19] based on the back-propagation from the stenosis classification decision and 2) anchor-based feature pyramid network (FPN) [20]. For FPN inputs, 1588 positioning boxes with a minimal size of 35 × 35 pixels were annotated by two independent expert cardiologists. The anchor-based model was trained with LearningRate=1e-4 over 500 epochs and using a pre-trained 2-CAT classifier as the backbone. Then FPN was built on the feature maps of pre-trained classification models. The same reader-annotated bounding boxes were also used for evaluation of the CAM-based localization technique. For the positioning training and 4-fold evaluation, 690 frames with >25% stenosis were used (Figure 1).

The performances of the two stenosis localization methods were assessed by the metrics of global-sensitivity, per-stenosis-sensitivity (Sens_s), per-stenosis-specificity (Spec_s), and mean square error (MSE). Global-sensitivity was defined as the recall of localization for the most significant stenosis in the images, which is similar to AR^(max=1) in COCO benchmark [21]. Sens_s and Spec_s were defined as the recall rate of all stenosis localizations in the images. MSE was assessed in 512 × 512 images for the CAM-based model and the anchor-based models. Due to the lower resolution, metrics for the CAM-based model were calculated with Intersection over Union (IoU) >0.2 in the CAM-based model whereas IoU>0.5 for the anchor-based model.

### Statistical Analysis

All the statistical evaluation was performed in Python (version 3.6; Python Software Foundation, Wilmington, Del; https://www.python.org). In this study, the calculation for diagnostic performance was based on a per-patient approach, including image-level severity classification. Accuracy, f1-score and Cohen’s Kappa were calculated for image-level stenosis classification; receiver operating characteristic (ROC) analysis and areas under the curves (AUC) were used to further evaluate the image-/artery-/patient-level diagnostic performance. Stenosis positioning was evaluated by sensitivity, specificity, and mean square error as described in the previous section. All metrics were computed using Scikit-learn, version 0.19.1. Continuous variables that were normally distributed were summarized and reported as means ± standard deviations.

## Results

### Patient Characteristics

The study participants’ characteristics are given in Table 2. A total of 230 individuals were included in our analysis. The median age was 62 years [IQR 55,69], 70% were men, 45% were white, 82% had hypertension, 71% had dyslipidemia, 16% were current smokers, and 27% had a high pretest probability of obstructive coronary artery disease.

**Table 2.**
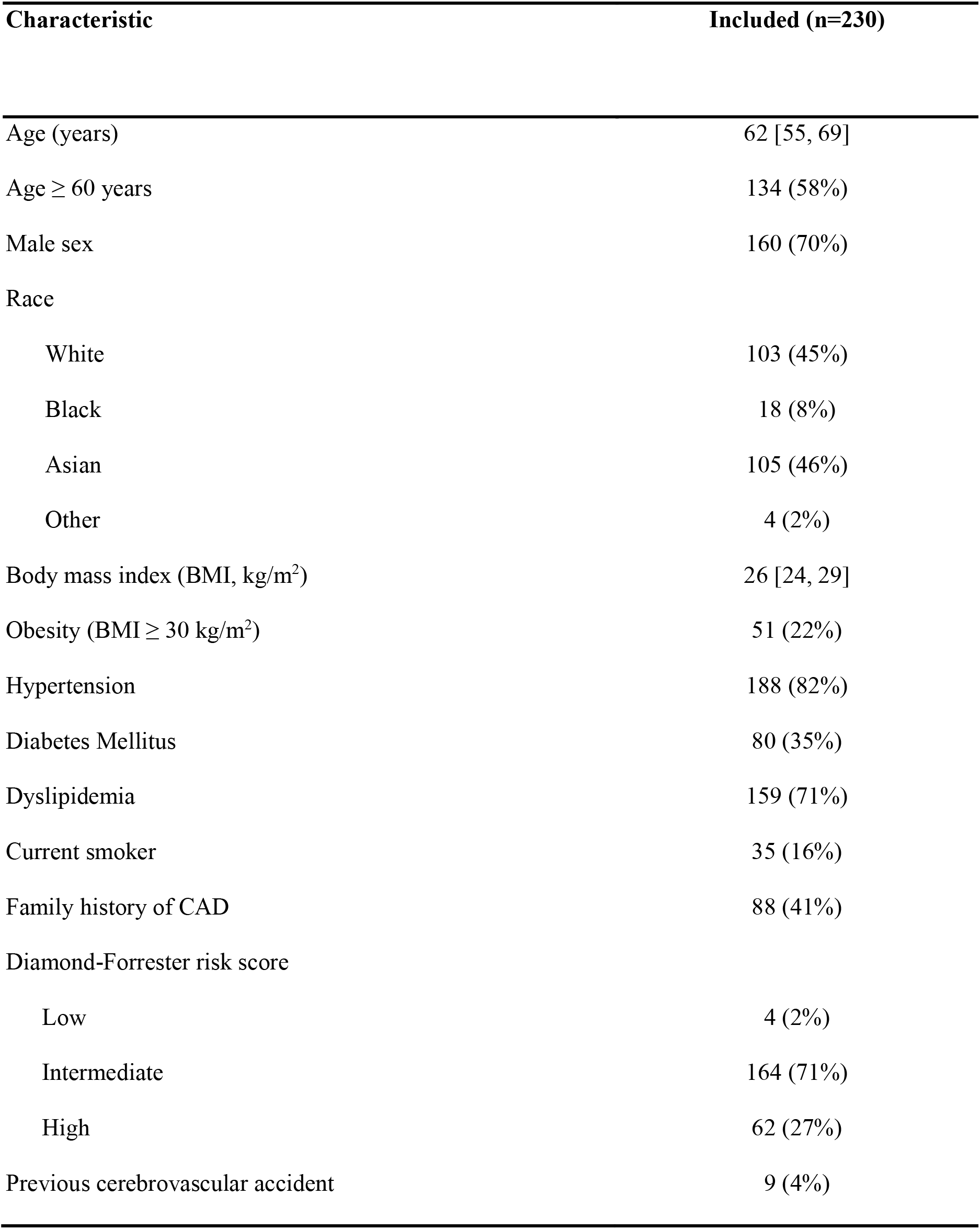
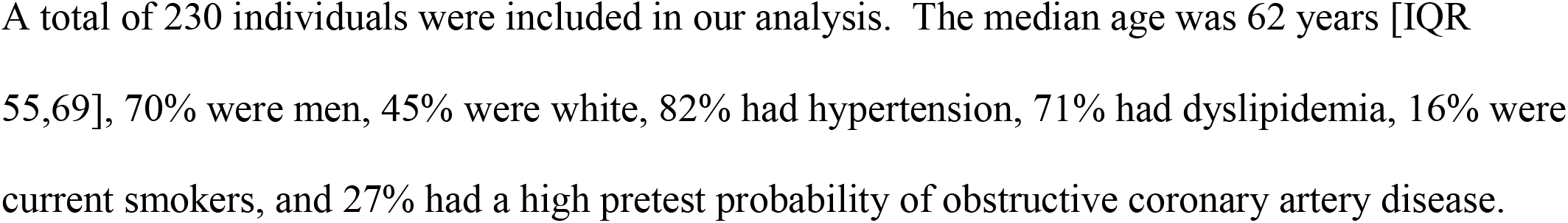
Clinical characteristics of the study participants.

### Candidate Frame Selection

The automatic model achieved a mean error of 2.05 and 2.27 in BCF and ECF detection respectively. The acceptance and error rates were 83% and 5.0%. A common feature of misclassified cases was a relatively short contrast duration in the video (typically <5 frames with adequate vessel-to-background contrast). The network did not adequately handle this type of condition because the training dataset had very few instances of short-duration contrasting frames.

### Stenosis Classification

The stenosis classification results in 3-CAT and 2-CAT with and without redundancy training models are summarized in Table 3 and Figure 3. In brief, the image-level classification performance was better in 2-CAT than 3-CAT for the LCA while not significantly different for the RCA. The redundancy training improved the AUC values for both 2-CAT and 3-CAT, as well as the accuracy, F1-score, and kappa score in 2-CAT. Based on the better performance in 2-CAT as well as our aim to pick-up non-significant stenosis, only 2-CAT evaluation was performed for artery-level (LCA and RCA) and patient-level classification. The accuracies were 0.83, 0.81, 0.85, the sensitivities were 0.94, 0.90, and 0.96 and AUCs were 0.87, 0.88, and 0.86 at the artery-level; LCA and RCA, and at the per-patient level respectively. A representative image illustrating the effect of the redundancy training is demonstrated in Figure 4 with visualization aided by a heatmap. The overfitting caused by background structures is markedly reduced, likely resulting in the improvement in classification performance.

**Table 3.**
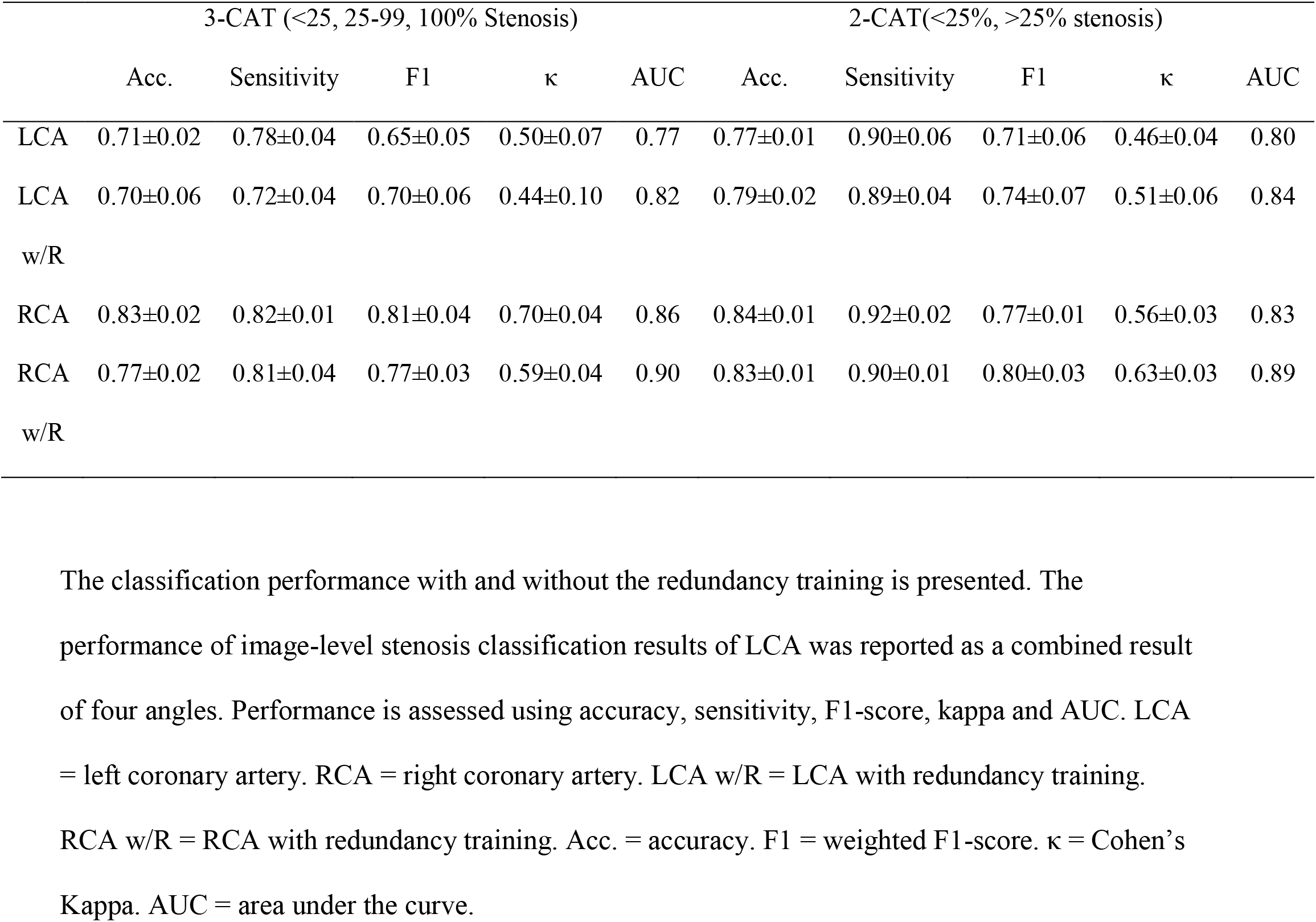
The image-level stenosis classification performance for the 2-category and 3-category severity levels.

**Figure 3.**
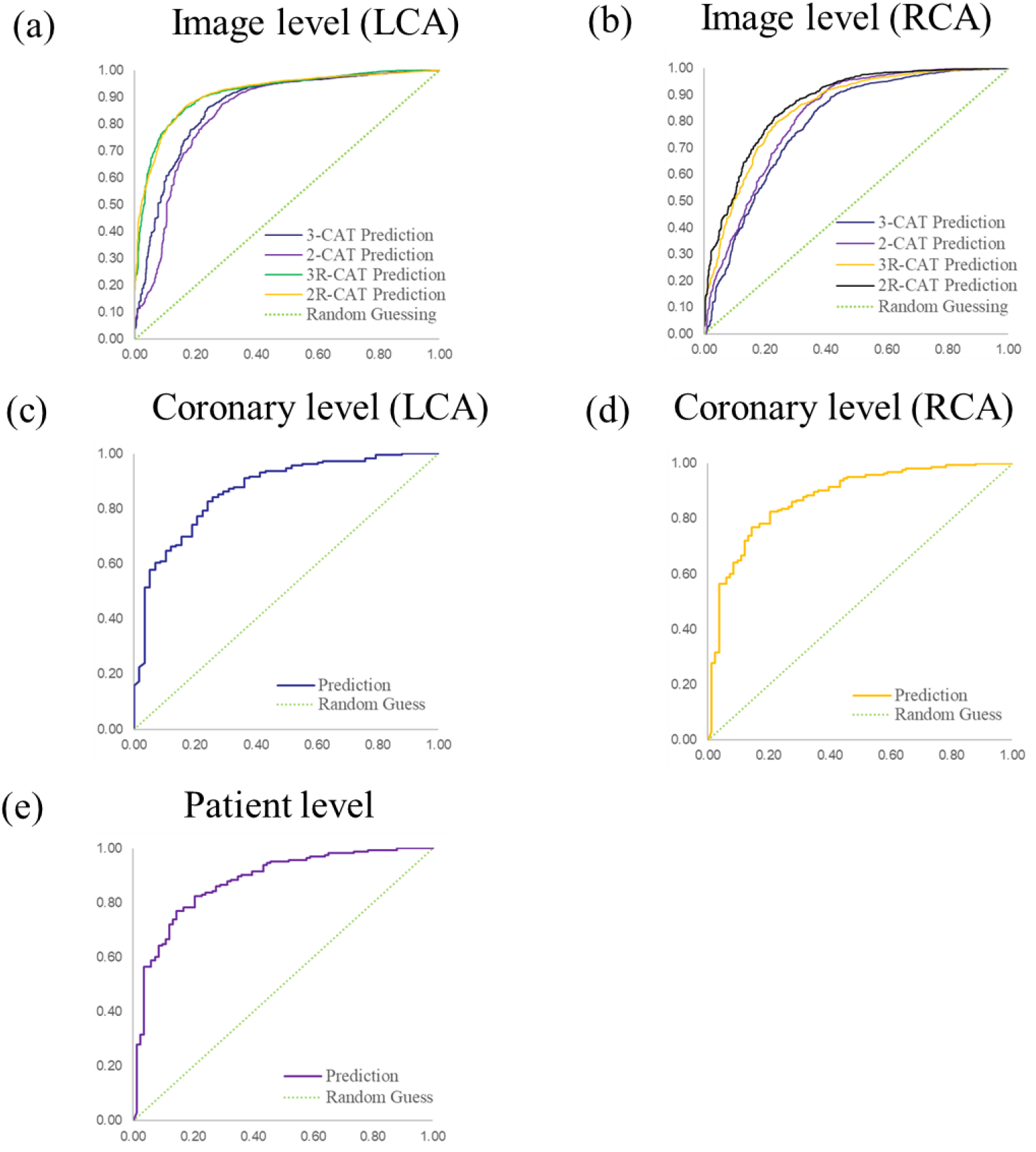
Performance of coronary stenosis classifications in image, coronary artery, and patient levels. (a) and (b): ROC curves of image level classification on 3-CAT and 2-CAT with and without redundancy training on LCA and RCA. (c) and (d): ROC curves of coronary artery level classification on LCA and RCA. (e): ROC curve of patient level classification. The AUC values are summarized in Table 3. RCA = right coronary artery. LCA = left coronary artery. AUC = area under the curve.

**Figure 4.**
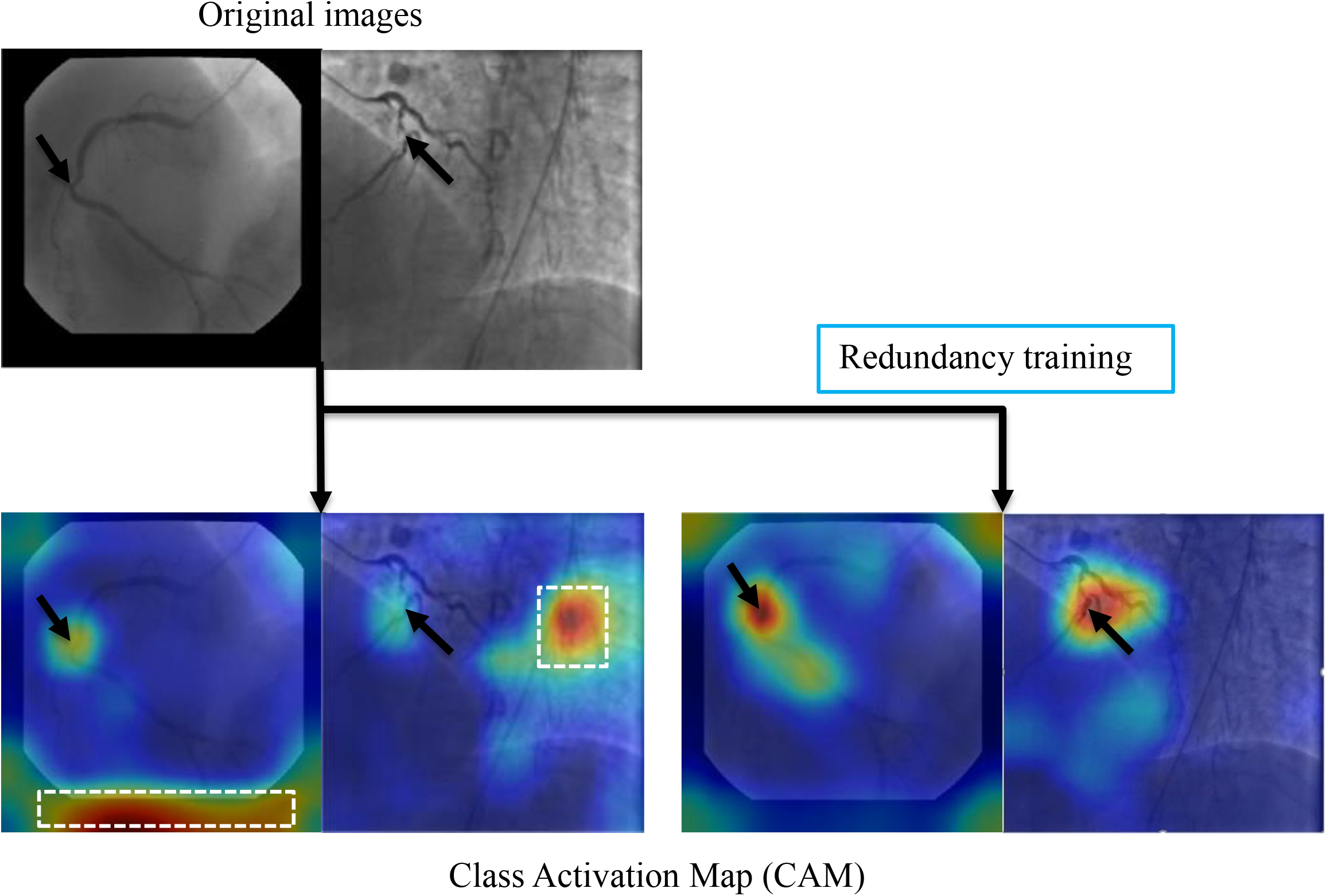
A representative image of the effect of the redundancy training demonstrated in a heatmap style. In the original training, the model had mid-to-high level attention on background regions. The redundancy training reduced the overfitting caused by background structures and improved the performance of stenosis classification.

### Stenosis Localization

Quantitative results were summarized in Table 4. In brief, the anchor-based FPN method showed better performance than the CAM-based method by all the metrics studied. Both the localization techniques performed better for RCA images than for LCA images. In both methods, Sensitivity was low due to the many annotations that highlighted small lesions that had ambiguous feature patterns in the arteries. Performance was also lower when there were multiple stenoses in distal coronary arteries or branches (see Figure 5 for illustration).

**Table 4.** Performance of the stenosis localization algorithms for the LCA and RCA.

|  | CAM-based |  |  |  | Anchor-based |  |  |  |
| --- | --- | --- | --- | --- | --- | --- | --- | --- |
|  | Global-sensitivity | Sens<sub>s</sub> | Spec<sub>s</sub> | MSE<br>(Deviation) | Global-sensitivity | Sens<sub>s</sub> | Spec<sub>s</sub> | MSE<br>(Deviation) |
| LCA | 0.59 | 0.25 | 0.43 | 103.3 (71.18) | 0.68 | 0.44 | 0.68 | 39.3 (40.00) |
| RCA | 0.61 | 0.17 | 0.51 | 79.5 (47.21) | 0.70 | 0.51 | 0.77 | 37.6 (51.63) |

Results are presented as the global sensitivity, sensitivity, specificity, and MSE for the two techniques presented – the CAM-based model and the anchor-based model. Global-sensitivity was defined as the sensitivity of one most severe stenosis localization per image. Due to the low resolution, metrics (Sens, Sens<sub>s</sub>, Spec<sub>s</sub>) for the CAM-based model were calculated with IoU > 0.2 whereas the metrics for the anchor-based model were calculated with IoU > 0.5. LCA = left coronary artery. RCA = right coronary artery. Sens<sub>s</sub> = per-stenosis sensitivity. Spec<sub>s</sub> = per-stenosis specificity. MSE = mean square error. CAM = class activation map. IoU = intersection over union.

**Figure 5.**
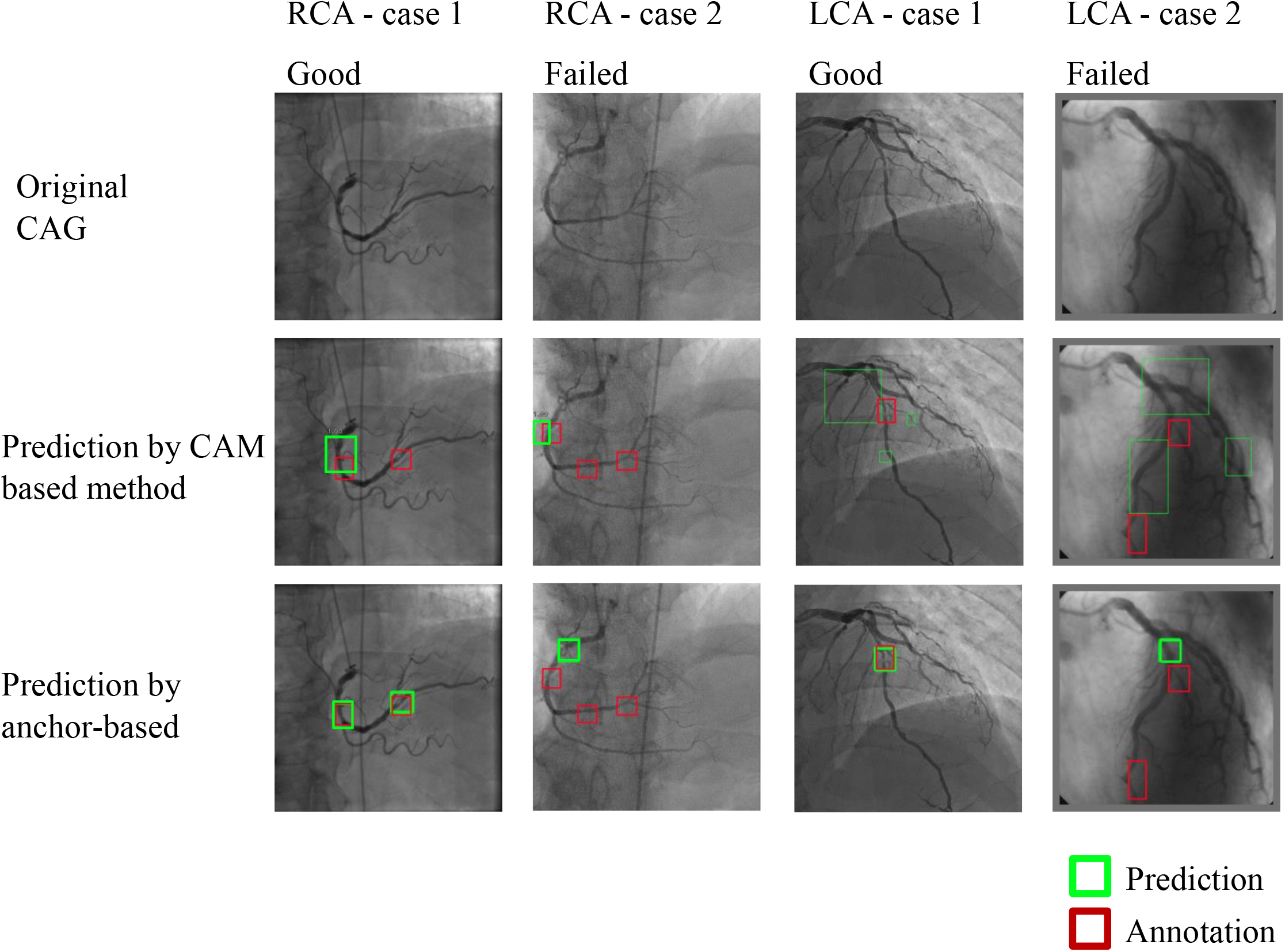
Representative images of stenosis position localization experiments. Predicted boxes from anchor-based model produced more accurate boxes when compared to the CAM-based model. Multiple stenoses in distal coronary arteries or branches were difficult for correct localization, which was the main reason for the failed cases.

## Discussion

The main findings from the present study are summarized as follows: 1) the fully-automatic, end-to-end workflow, which eliminated the vessel extraction and segmentation step for supervised-learning, was precise for stenosis classification on CAG images, achieving an accuracy of 0.85, a sensitivity of 0.96 and an AUC of 0.86 at the per-patient level. 2) Redundancy training improved the accuracy, F1-score, and the kappa score, and the AUC values for the image-level stenosis classification. 3) Stenosis localization was assessed by two methods: CAM-based and anchor-based models, with superior quantitative results for the anchor-based models.

End-to-end workflow is advantageous in reducing human interaction steps. In our proposed workflow, once applied to the CAG videos, the model automatically selects the optimal frames, performs stenosis classification and localizes stenosis positions, providing robust results at both the artery and patient levels. This workflow is advantageous in a large volume clinical setting or quality control purposes because the timely screening of many CAG videos to correctly identify cases with normal or mild stenoses can translate into improved productivity. The candidate frame selection performance presented here showed good results. This was better than what has previously been reported [2], likely due to the use of the bi-directional LSTM network in temporal information processing [22]. Additionally, by providing stenosis classification and localization, the reader/physician can verify the performance of the presented CNN framework and perform quantitative coronary angiography faster.

The stenosis classification results presented in our study are encouraging. We compared our stenosis classification performance with that of a previous CT study [23] with accuracies of 0.75/0.80 in 3-/2-CAT; and with three other vessel-based CAG studies [4], [5], [24] in 2-CAT with accuracies of 0.97/0.94/0.75 respectively. Therefore, our method was comparable and sometimes outperformed methods reported in previous studies. We attribute our favorable results to addressing different aspects of a typical CAG study such as multiple angle views, background frames and visually insignificant features of vessel stenoses through redundancy training to reduce overfitting in classification training.

Our study also explores a solution to the stenosis localization problem via an object detection framework. Two different stenosis localization methods of CAM and FPN were compared. FPN showed a better performance for stenosis positioning, even though additional annotations were necessary for training the algorithm. The CAM-based model has the strength of employing a simple derivation that uses stenosis classification as a backbone model.

Our study had a few important limitations. Training and evaluation were performed in the same cohort. A validation study using an external cohort is needed to accurately assess the performance of our techniques. Stenosis classification was simply categorized into three groups of <25% stenosis, 25 to 99% stenosis, and total occlusion for 3-CAT while <25% stenosis and 25 to 100% stenosis for 2-CAT. Our aim was to develop a tool that identifies normal and mild stenosis cases within a large cohort. In this regard, more granular categories for mild to moderate stenosis may be considered for different clinical or investigational purposes, such as detection of hemodynamically significant stenosis.

In conclusion, a fully-automatic end-to-end deep learning-based workflow which eliminates the vessel extraction and segmentation step was accomplished for accurate stenosis classification and localization on CAG images. This end-to-end approach may be useful in clinical settings with large patient volumes or for quality control of CAG procedures.

## Data Availability

The datasets used and/or analyzed during the current study are available from the corresponding author on reasonable request.

## Declarations

### Funding

This study was supported by Master Research Agreement 09-115 and Artificial Intelligence Health Information Exchange (AIHEX).

### Conflicts of interest/Competing interests

Dr. Joao A.C. Lima reports receipt of grant support from Canon Medical Systems. There are no conflicts of interest to declare for the remaining co-authors.

### Author contributions

Both Chao Cong and Yoko Kato are the first authors that equally contributed to this manuscript. Both of them worked on the data analysis, study design, and the manuscript drafting. All authors read and approved the final manuscript.

### Ethics approval and consent to participate

The original CORE320 study was approved by central and at 16 local institutional review boards (IRBs) which are listed below. Written informed consent was obtained from all participants as presented in the original CORE320 papers. (Clinical trial design paper by Vavere et al. Journal of Cardiovascular Computed Tomography 2011; 5: 370–381. Original study paper by Rochitte et al. European Heart Journal 2014; 35: 1120–30.) Given the retrospective and ancillary nature of the data, the current study is covered by the original CORE320 study IRB.

### List of the central and the 16 local institutional IRBs of the original CORE320 study

#### The central IRB

- Johns Hopkins School of Medicine (Study Chair: Joao AC Lima, MD MBA)

#### The local IRBs (16 centers)

1. United States (4 sites)
  - Johns Hopkins School of Medicine (PI: Joao A. C. Lima, MD)
  - National Heart Lung and Blood Institute (NHLBI) (PI: Andrew E. Arai, MD)
  - Beth Israel Deaconess Medical Center (PI: Roger Laham, MD)
  - Brigham and Women Hospital (PI: Frank J. Rybicki, MD)
2. Brazil (2 sites)
  - Albert Einstein Hospital (PI: Cesar Nomura, MD)
  - INCOR Heart Institute University-Sao Paulo (PI: Carlos E. Rochitte, MD, PhD)
3. Canada (1 site)
  - Toronto General Hospital (PI: Narinder Paul, MD)
4. Denmark (1 site)
  - Rigshospitalet - University of Copenhagen (PI: Klaus F. Kofoed, MD)
5. Germany (1 site)
  - Charite Humboldt University (PI: Marc Dewey, MD, PhD)
6. Japan (4 sites)
  - Iwate Medical University (PI: Kunihiro Yoshioka, MD)
  - Keio University (PI: Sachio Kuribyashi, MD)
  - St. Luke’s International Hospital (PI: Hiroyuki Niinuma, MD, PhD)
  - Mie University (PI: Hajime Sakuma, MD)
7. Netherlands (1 site)
  - Leiden University (PI: Joanne D. Schuijf, PhD)
8. Singapore (2 sites)
  - Mount Elizabeth Hospital (PI: John Hoe, MD)
  - National Heart Center (PI: Tan Swee Yaw, MD)

### Clinical trial registration

www.clinicaltrials.gov, NCT00934037

### Consent for publication

All subjects provided written informed consent including the publication of the data.

## Notes

### Clinical Trial

NCT00934037

### Author Declarations

The original CORE320 study was approved by central and at 16 local institutional review boards (IRBs) which are listed below. Written informed consent was obtained from all participants as presented in the original CORE320 papers. (Clinical trial design paper by Vavere et al. Journal of Cardiovascular Computed Tomography 2011; 5: 370-381. Original study paper by Rochitte et al. European Heart Journal 2014; 35: 1120-30.) Given the retrospective and ancillary nature of the data, the current study is covered by the original CORE320 study IRB. List of the central and the 16 local institutional IRBs of the original CORE320 study: The central IRB: Johns Hopkins School of Medicine (Study Chair: Joao AC Lima, MD MBA) The local IRBs (16 centers): 1. United States (4 sites) Johns Hopkins School of Medicine(PI: Joao A. C. Lima, MD) National Heart Lung and Blood Institute (NHLBI)(PI: Andrew E. Arai, MD) Beth Israel Deaconess Medical Center (PI: Roger Laham, MD) Brigham and Women Hospital (PI: Frank J. Rybicki, MD) 2. Brazil (2 sites) Albert Einstein Hospital (PI: Cesar Nomura, MD) INCOR Heart Institute University-Sao Paulo (PI: Carlos E. Rochitte, MD, PhD) 3. Canada (1 site) Toronto General Hospital (PI: Narinder Paul, MD) 4. Denmark (1 site) Rigshospitalet - University of Copenhagen (PI: Klaus F. Kofoed, MD) 5. Germany (1 site) Charite Humboldt University(PI: Marc Dewey, MD, PhD) 6. Japan (4 sites) Iwate Medical University (PI: Kunihiro Yoshioka, MD) Keio University (PI: Sachio Kuribyashi, MD) St. Luke's International Hospital (PI: Hiroyuki Niinuma, MD, PhD) Mie University (PI: Hajime Sakuma, MD) 7. Netherlands (1 site) Leiden University (PI: Joanne D. Schuijf, PhD) 8. Singapore (2 sites) Mount Elizabeth Hospital (PI: John Hoe, MD) National Heart Center(PI: Tan Swee Yaw, MD)

## References

[1] E. J. Benjamin et al., “Heart Disease and Stroke Statistics-2019 Update: A Report From the American Heart Association.,” Circulation, vol. 139, no. 10, pp. e56–e66, Mar. 2019.

[2] H. Ma, P. Ambrosini, and T. van Walsum, “Fast Prospective Detection of Contrast Inflow in X-ray Angiograms with Convolutional Neural Network and Recurrent Neural Network,” Dec. 2019.

[3] S. Moccia, E. De Momi, S. El Hadji, and L. S. Mattos, “Blood vessel segmentation algorithms -Review of methods, datasets and evaluation metrics.,” Comput. Methods Programs Biomed., vol. 158, pp. 71–91, May 2018.

[4] C. B. Compas, T. Syeda-Mahmood, P. McNeillie, and D. Beymer, “Automatic detection of coronary stenosis in X-ray angiography through spatio-temporal tracking,” 2014 IEEE 11th Int. Symp. Biomed. Imaging, ISBI 2014, pp. 1299–1302, 2014.

[5] T. Wan, H. Feng, C. Tong, D. Li, and Z. Qin, “Automated identification and grading of coronary artery stenoses with X-ray angiography,” Comput. Methods Programs Biomed., vol. 167, pp. 13–22, 2018.

[6] M. Fatemi, S. Mirhassani, and E. Ghasemi, “X-ray Angiographic images using contour processing of segmented Heart Vessels based on Hessian Vesselness Filter and Wavelet Based Image Fusion,” Int. J. Comput. Appl., vol. 36, no. 9, pp. 27–33, 2011.

[7] A. Hernandez-Vela et al., “Accurate coronary centerline extraction, caliber estimation, and catheter detection in angiographies,” IEEE Trans. Inf. Technol. Biomed., vol. 16, no. 6, pp. 1332–1340, 2012.

[8] J. Brieva, M. Gálvez, and C. Toumoulin, “Coronary extraction and stenosis quantification in X-Ray angiographic imaging,” Annu. Int. Conf. IEEE Eng. Med. Biol. -Proc., vol. 26 III, no. 1, pp. 1714–1717, 2004.

[9] S. Yang et al., “Deep learning segmentation of major vessels in X-ray coronary angiography,” Sci. Rep., vol. 9, no. 1, pp. 1–11, 2019.

[10] E. Nasr-Esfahani et al., “Vessel extraction in X-ray angiograms using deep learning,” Proc. Annu. Int. Conf. IEEE Eng. Med. Biol. Soc. EMBS, vol. 2016-Octob, pp. 643–646, 2016.

[11] E. Nasr-Esfahani et al., “Segmentation of vessels in angiograms using convolutional neural networks,” Biomed. Signal Process. Control, vol. 40, pp. 240–251, 2018.

[12] K. Jo, J. Kweon, Y. H. Kim, and J. Choi, “Segmentation of the Main Vessel of the Left Anterior Descending Artery Using Selective Feature Mapping in Coronary Angiography,” IEEE Access, vol. 7, pp. 919–930, 2019.

[13] A. L. Vavere et al., “Diagnostic performance of combined noninvasive coronary angiography and myocardial perfusion imaging using 320 row detector computed tomography: Design and implementation of the CORE320 multicenter, multinational diagnostic study,” J. Cardiovasc. Comput. Tomogr., vol. 5, no. 6, pp. 370–381, 2011.

[14] C. E. Rochitte et al., “Computed tomography angiography and perfusion to assess coronary artery stenosis causing perfusion defects by single photon emission computed tomography: the CORE320 study.,” Eur. Heart J., vol. 35, no. 17, pp. 1120–30, May 2014.

[15] C. Szegedy, V. Vanhoucke, S. Ioffe, J. Shlens, and Z. Wojna, “Rethinking the Inception Architecture for Computer Vision,” Proc. IEEE Comput. Soc. Conf. Comput. Vis. Pattern Recognit., vol. 2016-Decem, pp. 2818–2826, 2016.

[16] J. A. Garcia et al., “Determination of optimal viewing regions for X-ray coronary angiography based on a quantitative analysis of 3D reconstructed models.,” Int. J. Cardiovasc. Imaging, vol. 25, no. 5, pp. 455–62, Jun. 2009.

[17] J. Wang, J.-M. Wei, Z. Yang, and S.-Q. Wang, “Feature Selection by Maximizing Independent Classification Information,” IEEE Trans. Knowl. Data Eng., vol. 29, no. 4, pp. 828–841, Apr. 2017.

[18] E. L. Alderman and M. Stadius, “The angiographic definitions of the bypass angioplasty revascularization investigation,” Coron. Artery Dis., vol. 3, no. 12, pp. 1189–1207, 1992.

[19] B. Zhou, A. Khosla, A. Lapedriza, A. Oliva, and A. Torralba, “Learning Deep Features for Discriminative Localization,” Acad. Manag. Proc., vol. 2004, no. 1, pp. M1–M6, Dec. 2015.

[20] T.-Y. Lin, P. Dollár, R. Girshick, K. He, B. Hariharan, and S. Belongie, “Feature Pyramid Networks for Object Detection,” Proc. - Int. Conf. Tools with Artif. Intell. ICTAI, vol. 2019-Novem, pp. 1702–1707, Dec. 2016.

[21] T.-Y. Lin et al., “Microsoft COCO: Common Objects in Context,” Lect. Notes Comput. Sci. (including Subser. Lect. Notes Artif. Intell. Lect. Notes Bioinformatics), vol. 8693 LNCS, no. PART 5, pp. 740–755, May 2014.

[22] A. Graves and J. Schmidhuber, “Framewise phoneme classification with bidirectional LSTM and other neural network architectures.,” Neural Netw., vol. 18, no. 5–6, pp. 602–10, 2005.

[23] M. Zreik, R. W. Van Hamersvelt, J. M. Wolterink, T. Leiner, M. A. Viergever, and I. Išgum, “A Recurrent CNN for Automatic Detection and Classification of Coronary Artery Plaque and Stenosis in Coronary CT Angiography,” IEEE Trans. Med. Imaging, vol. 38, no. 7, pp. 1588–1598, 2019.

[24] B. Au et al., “Automated Characterization of Stenosis in Invasive Coronary Angiography Images with Convolutional Neural Networks,” arXiv Comput. Vis. Pattern Recognit., Jul. 2018.

